# Technology Based Challenges of Informal Clinical Communication in an Australian tertiary referral hospital – A mixed methods assessment of The Need for Change

**DOI:** 10.1101/2024.06.26.24308798

**Authors:** Graeme K Hart, Nicole Hosking, Julia G Todd, Lorelle Martin

**Author notes:** **Corresponding Author:** Assoc Prof Graeme K Hart MB BS, FANZCA, FCICM, FAIDH.

## Abstract

**Background:** Effective communication is critical for safe, efficient clinical practice. Communication failures result in errors, misdiagnosis, inappropriate treatment and poor care. Communication errors also contribute to sentinel events and are an underlying factor in healthcare system complaints.

Formal Clinical Communication (FCC) tools, such as ISBAR, improve patient outcomes. Governance of FCC is increasingly based around Electronic Medical Record (EMR), however much Informal clinical communication (ICC) occurs outside of the EMR.

ICC involves disparate platforms including pagers, Short Message Service (SMS) texts, encrypted messaging apps, phones and local radio networks (eg Vocera). Documentation of ICC in the clinical record is low quality and not easily or routinely audited. ICC based on personal identities relies on accurate rosters, switchboard staff or secondary lists. Significant inefficiency and potential clinical risk can occur if the correct role to communicate with cannot be contacted quickly and easily.

**Local Problem:** In 2019, Austin Health performed a clinical governance assessment of ICC processes against National Standards for FCC. Further investigation and process mapping of ICC occurred in 2020. This indicated a paucity of relevant policy and procedures to govern ICC practices, with highly variable and overly complex processes.

**Aims:**

1. To document the technology used in informal communication between clinical and / or administrative staff.
2. To document the self-perceived impact on staff of current communications methods.
3. To document the self-perceived potential efficiency and safety impact of current communications methods.
4. To identify key factors for consideration in organisational informal communication improvement.

**Method:** Multi-disciplinary on-line staff cross-sectional survey using Microsoft Forms. The survey sought to confirm the range of informal communication methods in use and identify respondents’ perceptions of current multimodal communication technology issues and their inherent risks.

**Results:** 115 self-selected clinical and administrative staff completed the survey. Multiple communication channels are used. Respondents noted high levels of frustration, delay, interruption and inefficiency. Desired communication improvements and use considerations were identified. Survey findings validated a prior clinical governance assessment of the existing ICC framework, and the need for technology reform.

**Conclusions:** There are gaps in governance standards for ICC, both locally and at a broader level. Sequential additions to technology platforms have created a high-risk communications environment. Staff perceptions of inefficiency, delay, frustration and a high level of patient safety risk were consistent across professions. This work informed the subsequent development of an enterprise platform dedicated to informal clinical communication.

**Key Message Summary Box:**

- **What is already known on this topic** – Poor Communication practice contributes to preventable errors or adverse events in patient care. Use of structured handover tools such as ISBAR provide a framework to improve Formal Clinical Communication. Current multi-modal, communication technologies for Informal Clinical Communication are interruptive, inefficient, compound staff frustration and create potential for patient harm.
- **What this study adds** – We describe staff impressions of frustration, time wasting and potential for patient harm with existing multi-modal communications technologies together with a framework for informal communication policy enhancement.
- **How this study might affect research, practice or policy** – These findings provide a call for governance standards for informal clinical communication. We highlight the need for rationalisation of multi-modal communications technologies to reduce communication complexity and identify some key functional requirements for new technologies.

## Introduction

### Problem Description

Poor communication or communication failures remain a leading cause of harm and poor patient experience within the hospital setting. Communication is a contributing factor in 70% of adverse events and has been identified as the most attributable root cause within 65% of sentinel patient events (1) (2).

Formal Clinical Communication (FCC) processes, such as structured clinical handover are promoted by Safety and Quality authorities and are key accreditation assessments in Australia and internationally. Use of a structured communication tool such as SBAR and ISBAR, can significantly improve the quality and safety of clinical handover communications (3). Governance regarding Informal Clinical Communication (ICC) is much less represented within accreditation requirements and published research.

Following the revision of the Australian ACSQHC Version 2 accreditation requirements (4), a significant gap in Austin Health’s governance of clinical communication practice was identified. There appeared to be paucity of standards and process metrics for ICCs in the accreditation or governance literature.

A Gap Analysis was conducted whereby ICC processes were assessed against ACHS clinical communication governance criteria (ACSQHC standard 6 - 6.01, 6.02 and 6.04) (4), with comparison of formal (FCC) vs Informal Communications (ICC). (Figure 1). Whilst clinical handover (eg. shift to shift handover, ward rounds) had longstanding high levels of organisational governance and compliance with expected content, ‘informal’ communication was lacking in policy, procedure, structured tools and the ability to monitor and assess communication performance.

**Figure 1.**
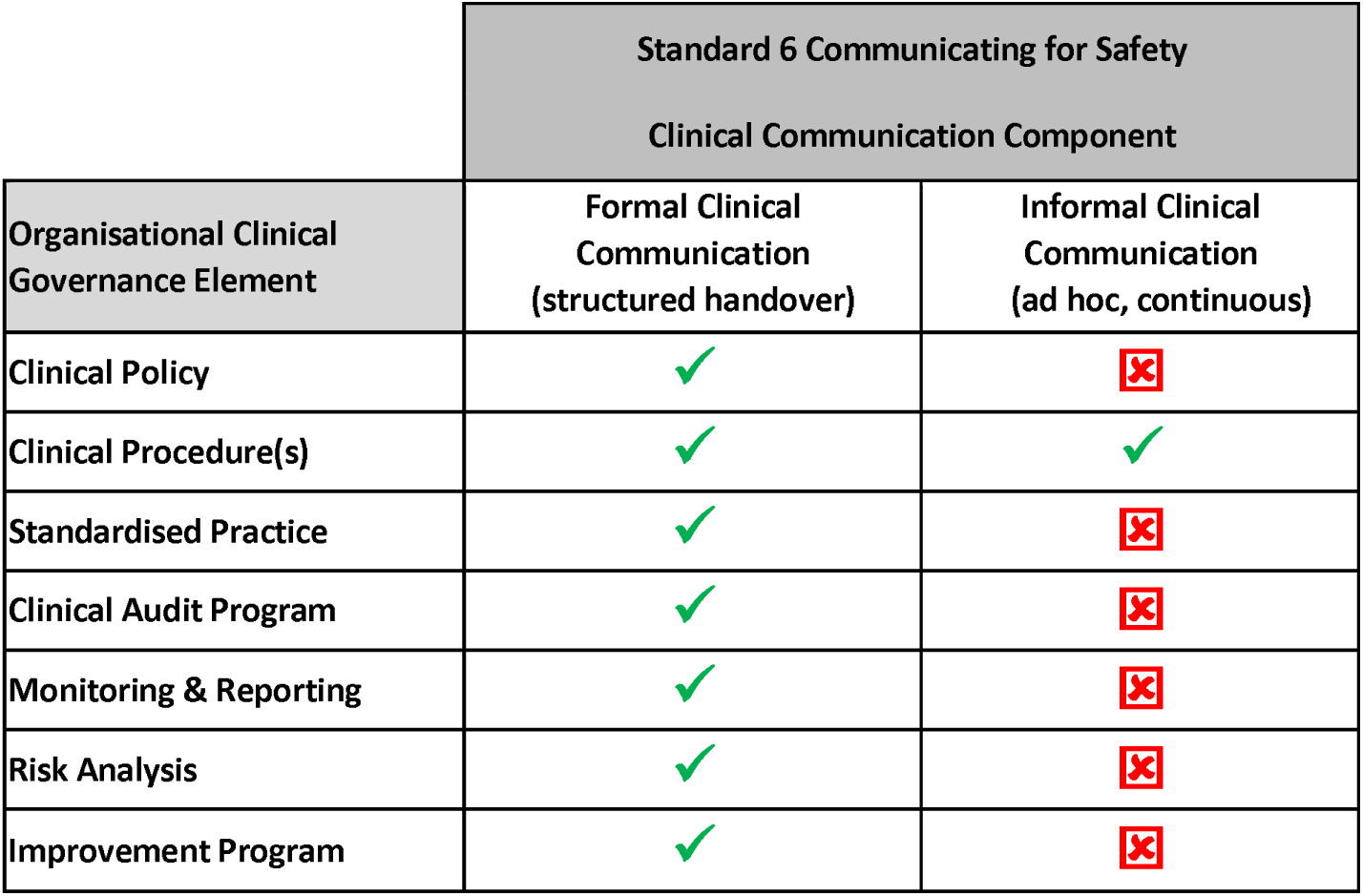
Comparison of explicit governance requirements for Informal Clinical Communication compared to Formal Clinical Communication in the Australian National Standards(4)

Based on this assessment, further investigation was performed to map existing informal communication processes in detail. The majority of communications occurring outside of structured clinical handover were ad hoc, inter-team, and role-based in nature. When mapped across the organisation, inter-team, role-based clinical communication practice was discovered to be an overly complex, highly variable system with two major issues identified (Figure 2). Firstly, it was difficult for front-line clinicians to identify who was performing a specific clinical role at a given point in time. Secondly, once a person was identified, there were multiple different communication methods available and individual receiver preferences often dictated the method used. Such preferences may be known at switchboard but are not generally visible.

**Figure 2.**
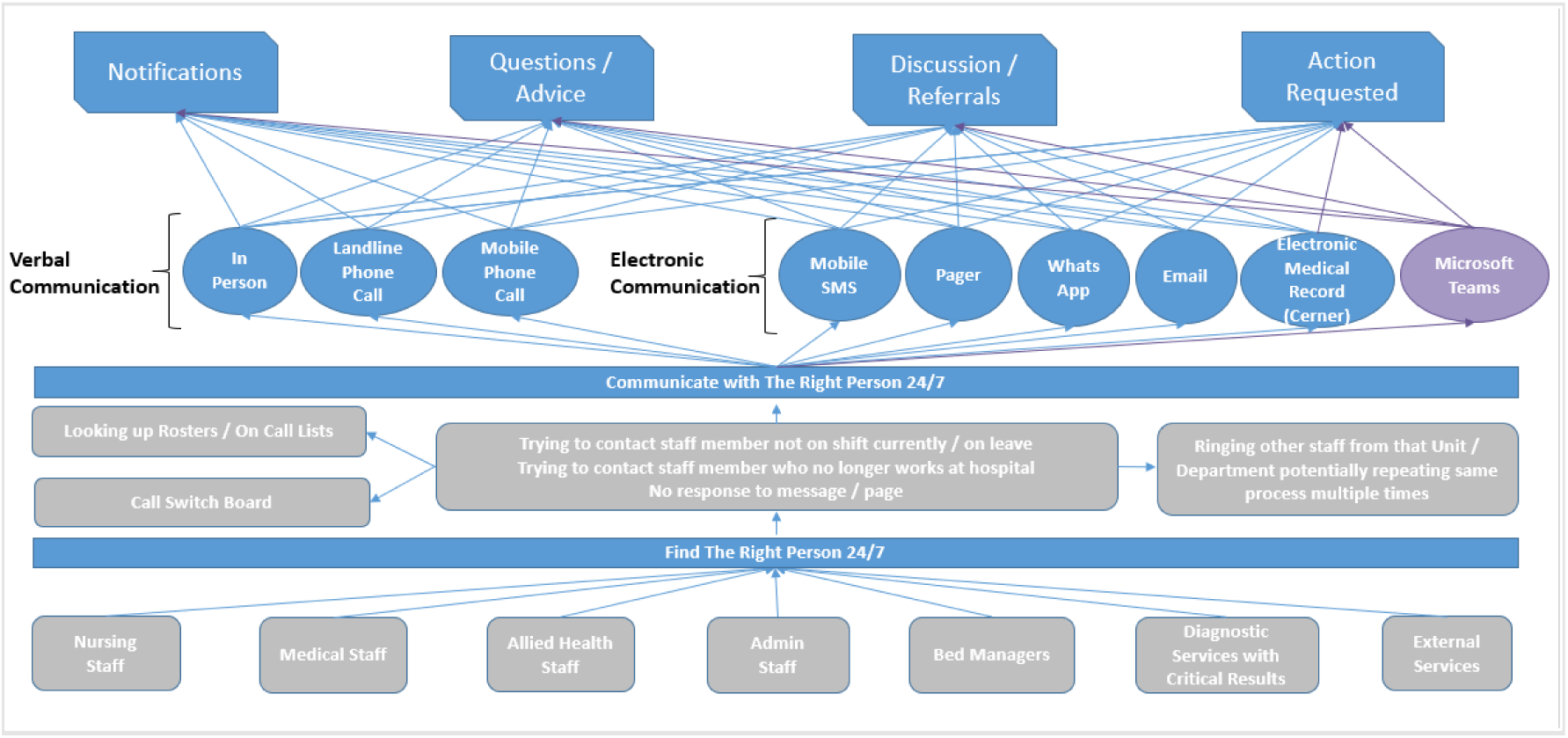
Patterns and technologies supporting functional Communications requirements observed during Health Service wide Communications Governance Review.

**Figure 3:**
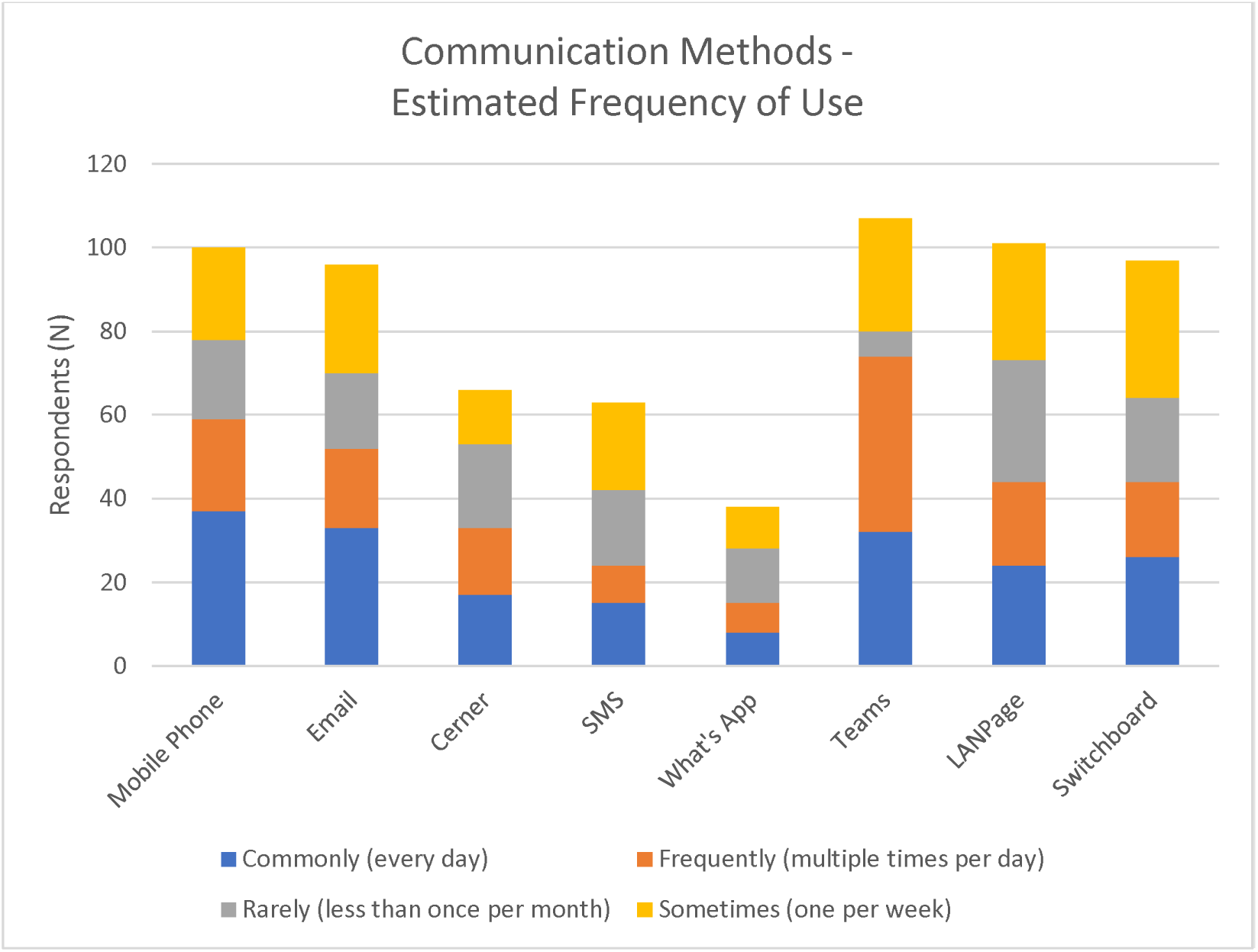
Respondent reported Estimated Frequency of Use of communications technologies.

Investigations into preventable patient harm recognised decades ago that the care environment is a major factor in sub-optimal human performance (5). Communications technologies impact on the practical processes of everyday communication but remain a relatively poorly studied component of the research. Hospitals have historically adopted new communication platforms over time, with little consideration for the need to rationalise or remove redundant technology. These disparate communication methods lead to inefficiencies and variations in practice. Comparison of 5 hospitals demonstrated significant difference in the benefits and unintended consequences between paging, Short Message Service (SMS), phone and computer-based systems. All created interrupted workflows and potential for patient risk and none offered solutions for all staff requirements (6).

Our process mapping indicated that communication technologies in use included paging, SMS, telephone calls to and from mobile (personal and hospital owned) and fixed telephone numbers, email, a variety of unsanctioned communication platforms such as WhatsApp and, more recently, Microsoft (MS) Teams. MS TEAMS had been introduced to general health service use in 2020 during the initial COVID-19 pandemic in Victoria, but was not initially configured as a clinical communication tool.

Some hospital issued pagers and phones were handed off between staff performing specific roles for which those devices were provided. However, most of these communication technologies, are reliant on the parties knowing the name and contact details of the individual with whom they need to communicate. This role may be filled by multiple staff over a shift cycle. Each of these technologies offered benefits at the time of introduction, but no single platform could satisfy the requirements identified in the proposed ICC standard. The majority demonstrated high-variation, poor documentation, poor visibility and accessibility, one-directional (asynchronous) communication and disruptive impact.

Improvements to the governance and day to day performance of ICC was deemed a ‘Communicating for Safety’ (Standard 6) (4) priority. To help inform a revision of the organisational role-based clinical communication framework and supporting technology, a survey of clinical staff was performed to validate the process map findings and understand the perceived impacts on clinician experience.

## Aims

### Setting

Austin Health is a 1000 bed, 3 campus, tertiary/quaternary health service in Melbourne Australia. Clinical services include general and specialist surgery and medical services, Emergency Department, Intensive Care, Liver, Kidney and Bone Marrow transplant, cardiothoracic surgery, oncology, mental health.

## Methods

A cross-sectional staff survey was conducted in 2021 over a 3-month period during the pre and early introductory phase of the pilot project. The key concepts identified in the Gap Analysis were incorporated into the Survey tool (Appendix 1 – online resources) seeking to estimate the frequency and nature of the technology platforms used for ICC. The survey tool asked users for estimated frequency of use, impact of technology and work practice required to identify the correct target person / role and the perceived impact of interruptions on patient safety and the respondent’s work. Survey tools had some role (nursing, medical, allied health) specific fields for role delineation but were otherwise identical across professions. Patients and Consumers were not involved in this study.

Multi-disciplinary clinical and non-clinical staff in the pilot clinical units were invited by email and staff notices via Microsoft Teams, email and paper notices to complete an electronic survey. The survey used a voluntary on-line MS FORMS based application. Staff roles, but not personal respondent identifiers were retained. Only aggregated group summaries were used.

The survey tool was developed by the senior project staff and was structured to identify user perceptions relating to their use of ICC technologies and the implications this had on their day to day professional tasks, patient safety and efficiency. It contained 15 questions asking respondents to select a standardised response or rate an item on a 5 point likert scale. The survey is provided in Appendix 1 on line resources.

Survey data was collated and analysed using MS Excel and Data was downloaded from MS FORMS as role specific Excel spreadsheets which were amalgamated into a single table. Pivot charts and Graphs were created. Responses were aggregated by groupings with more than 5 respondents per cell to protect respondent confidentiality. Reporting methodology was informed by the CROSS criteria(7)

### Ethics

This component of the project was undertaken as a QI project with Project number HREC 87684/Austin 2022.

## Results

### Staff survey

115 respondents completed the survey. 24 departments and 20 role categories were represented.

Respondent Demographics were broad and included Medical, Nursing, Allied Health, Laboratory, Radiology and Administrative staff and are shown in Table 1.

**Table 1.**

| 1. Respondent Department |  | Respondent Roles |  |
| --- | --- | --- | --- |
| Department | Respondent Number |  | Respondent Number |
| Ambulatory Care Centre | 1 | ANUM | 6 |
| Anaesthesia | 3 | Bedside Nurse | 14 |
| Cardiac Ward 5 East | 1 | NUM Liaison / Coordinator | 5 |
| Continuing Care | 1 | Dietitian | 12 |
| Diagnostics | 2 | Allied Health Practitioner | 23 |
| Emergency Department | 22 | Radiographer | 7 |
| General Medicine | 5 | Intern/HMO | 6 |
| Gynaecology | 1 | Registrar/Fellow | 12 |
| Intensive Care | 9 | Consultant Medical | 34 |
| Mental Health | 2 |  |  |
| Molecular Imaging and Therapy | 1 |  |  |
| Nutrition & Dietetics | 13 |  |  |
| Occupational Therapy | 12 |  |  |
| Oncology / Palliative Care | 3 |  |  |
| Orthotics & Prosthetics | 2 |  |  |
| Paediatrics | 1 |  |  |
| Physiotherapy | 2 |  |  |
| Radiology | 16 |  |  |
| Renal | 2 |  |  |
| Speciality Medicine | 6 |  |  |
| Speech Pathology | 4 |  |  |
| Spinal | 1 |  |  |
| Sub-acute | 1 |  |  |
| Surgery | 4 |  |  |
| **Total** | **115** |  | **115** |

**Table 2.**
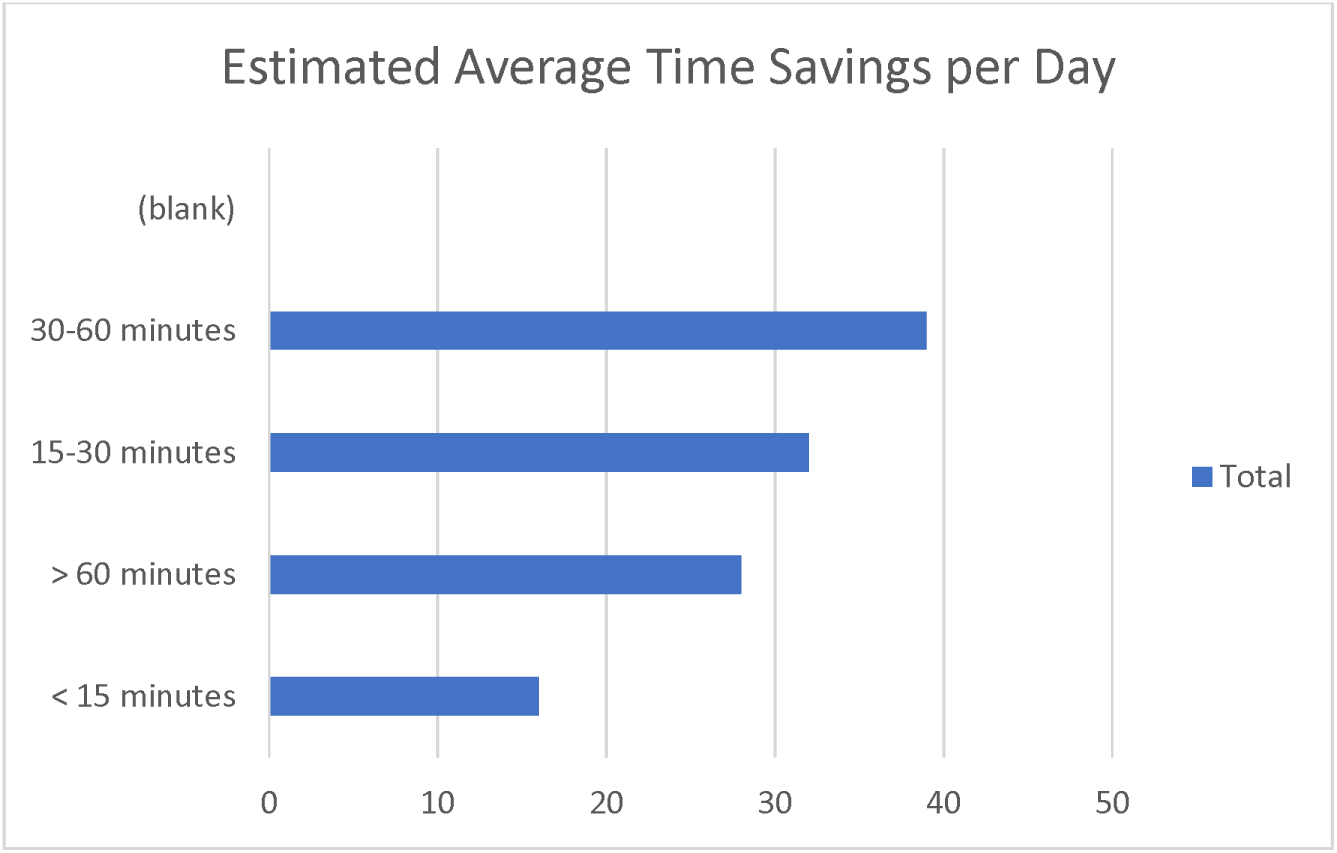

### Utilisation patterns of existing technology

Significant variation in the use of clinical communication platforms was reported. Some roles had no access to clinical systems such as CERNER EMR, however all other options were available to all respondents. TEAMS had already been implemented across the organisation and was available for general administrative use. It was already being used informally for some clinical communications by many staff. SMS messaging was used by approximately one third of respondents, as was the use of encrypted messaging services such as WhatsApp, despite the latter not being a hospital endorsed communication platform. Hospital switchboard, LAN Page and mobile phones played a very significant role for staff attempting to reach the clinician they needed to contact. Asynchronous messaging communication via LAN Page was indicated to be used frequently (multiple times per day) or commonly (everyday) by 38.3% of respondents.

### Efficiency and Effectiveness of current methods

#### Find First Time

Many respondents described difficulty in contacting relevant staff on the first attempt. 55 of 115 (47.8%) described inability to contact at first attempt more than 50% of the time. This issue may explain the continuing use of switchboard services to aid in finding the right person, as switchboard have access to the most accurate staffing rosters and are also able to re-direct calls to covering clinicians as required.

#### Potential time savings

Respondents believed they would save significant amounts of time each day if they were able to contact the right person on the first attempt. 39/115 (33.9%) anticipated they would save between 30 and 60 minutes per day and 28/115 (24.3%) estimated time savings of over 1 hour per day. This could have significant follow-on implications for patient flow activities, if more timely discharge planning and additional clinic appointments could be achieved because of improved clinical communication efficiencies.

#### Response Times

Respondents estimated Timely (within 30 minutes) or Reasonable (within 1 hour) response times 89/115 (77.4%) of the time.

#### Impact of time of Day

52/115 Respondents did not believe that there was a difference in terms of response times between “In hours” or “after hours” (in this case loosely defined as AM or PM). However, 32/115 described weekends and 18 /115 described After Hours as being a significant factor to the ability to contact the right person. The 2 cohorts reporting “out of hours impacts” were bedside nurses and consultant medical staff.

### Clinical Disruption by Interruption

56 /115 respondents described significant disruptions to clinical work related to incoming communications calls or messages. 19/115 (16.5%) reported between 10-20 interruptions per shift and 11 /115 (9.6%) reported more than 20 interruptions per shift.

#### Potential impacts of Current Communication Methods were listed

Respondents rated their level of agreement for each. 90/115 (78%) agreed with respect to impacts on keeping track of communication and related actions, 93/115 (81%) agreed with missing communications, 86/115 (75%) agreed with No Response to Communication, 85/115 (75%) agreed with No Documentation, 52/115 (45%) agreed with 3 Points of Identification not being used. 28/115 (28%) agreed that Adverse Events were more likely, and /115 (57%) agreed that there was Interruption to clinical tasks.

83% of respondents agreed that they did not consistently receive all required information more than 80% of the time. Missing important information included Patient identifiers, 24/115 (21%), Call back / return information 23/115 (20%), Identification of initiator 20/115(19%), Nature of problem 25/115 (22%), acknowledgement of request 8/115 (7%). 20% of respondents 23/115 reported being the incorrect recipient of a clinical communication at least once per day.

#### Documentation of the Communication

Only 20 of 115 (17.4%) respondents believed that the current communications tools provided adequate documentation of the interaction. This finding is consistent with prior governance concerns regarding poor documentation discovered during audits of clinical incidents.

## Discussion

The imperative to improve clinical communication is well recognised by Australian and international health and quality of care agencies. FCC methods such as clinical handover are widely promoted by Safety and Quality authorities, and Accreditation systems are designed to improve safety in the transfer of critical information (4, 8) Structured handover communication tools such as SBAR and ISBAR content models, provide a clear framework for clinical communication to reduce the likelihood of information gaps in specific circumstances and help ensure key information is included in the transfer of information between clinicians (9, 10).

ICC involves high-volume, ad-hoc, and unstructured communication processes such as in-person conversations, pages, phone calls, role based wireless communications devices (eg Vocera (11)) and electronic messaging via personal or hospital provided smart devices across the 24hr clinical care environment (12). Australian accreditation requirements currently contain limited assessment of ICC processes. Accreditation schemes also do not address communication at a technology level, despite years of clinical communication research demonstrating both negative and positive impact of specific clinical communication methods on the quality and safety of clinical communication practice (13), (14), (15, 16) (17).

Sociotechnical frameworks for ICT implementations have been proposed (18) and recommendations for the safe use of technology identified (19) although these largely focus on EMR based communications and design. There have been far fewer publications describing the impact of the communication technology on the safe delivery of clinical care, workforce satisfaction, clinician frustration and burnout. Those publications which do exist, highlight differences between synchronous and asynchronous tools, the differences in use cases, clinician preference for modalities and the high burden of use. Synchronous communications provide most information density and interaction but are the most interruptive (20). While there are many publications on ‘clinical burnout’ related to the EMR, and some on a single communication method, there are limited publications regarding the impact of multi-modal communication technology (21).

Through a detailed governance assessment of clinical communication practice, Austin Health identified high variability of practice for these predominantly role-based ICCs occurring outside of formal structured handover processes. This assessment highlighted four major classifications of informal multi-disciplinary communication – Notifications, Questions and Advice, Referrals - Discussion and Action Requests. Similar classifications of clinical communication have been previously described in an analysis of junior medical staff task management activity (22). Our assessment findings also indicated the use of a variety of technology platforms in use including paging, short message service (SMS), fixed line telephone calls, mobile phone calls, email, a variety of unsanctioned communication platforms such as WhatsApp and, more recently, Microsoft Teams.

Through the staff survey conducted in 2022, we confirmed the range of clinical communication technologies currently in use and identified a range of issues associated with this complex multi-modality communication system. Survey results revealed paging, phone calls and Microsoft Teams as the dominant forms of communication technology in use. Issues identified with existing communication technologies related to communication timeliness, communication quality and disruption to clinical care delivery. The patient safety risks and potential impacts attributable to the issues identified, are in alignment with existing communication research evidence, particularly those relating to the use of paging and phone communication technology (20) (23),(24, 25) (26)

From a communication timeliness perspective, 44% of respondents reported inability to make contact at first attempt more than 50% of the time. Survey results indicated approximately 18% of all communications receive poor to no response at all. 58.2% of survey respondents believed they would save significant amounts of time each day (>30min) if they were able to contact the right clinician on first attempt. 38% of respondents use switchboard commonly or frequently, suggesting that the identification of correct person to, or modality for, contact may be difficult. These findings would be consistent with the challenges identifying correct personnel described in Emergency Departments and supports the concept of Role-based Communications (27).

Despite advancements in electronic communication tools, the ongoing use of text (alphanumeric) paging for clinical care delivery remained considerable (38% respondents). The reported impact of the use of pagers was in alignment with previous studies that have highlighted the inefficiency associated with poor visibility and inaccuracy of pager allocation processes. Significant limitations in real-time communication occur using paging technology in an acute care setting (13) (14). Use of pagers not only raised concern from a communication timeliness perspective, but also from a communication quality perspective given key information is often omitted.

Only 11% of survey respondents believed that they received all required information during clinical communication more than 80% of the time. The majority of survey respondents (82%) identified missing communication as having a potential negative impact on workflows and efficiencies of clinical care. Lack of patient identifiers, call back / return information, sender information, and description of the nature of the problem were cited as issues in the quality of electronic communication performed. Text paging has been found to lack clarity, contain poor messaging structure, and have high variability of messaging content (14). Austin Health staff reports were in alignment with these findings.

The use of paging communication also requires consideration with relation to clinical service disruption. Cognitive overload is common in large, complex healthcare settings and pager-based interruptions have been identified as one cause (23). The number and variety of interruptive communications that occur contributes to cognitive overload, frustration, and error. A multihospital incident reporting system showed 220 potentially harmful incidents related to “interruption” and “distraction”, almost 10% of these were technology related. Phone calls have also been identified as a common interruption to the delivery of clinical care (21, 28), (29), with interruptions in the performance of clinical tasks an established contributing factor in specific adverse patient events including medication errors and procedure failures (30), (31), (24).

Phone calls via mobile (51.3% respondents) and fixed lines (42.6% respondents) were identified as a predominant form of communication within the existing Austin Health clinical communication framework. 48.7% of respondents described significant interruption to clinical work related to phone calls. 16.5% of respondents reported 10-20 interruptions per shift and 9.6% of respondents reported more than 20 interruptions per shift. This data is consistent with international findings (29), although our self-reported interruptions, despite being significant in frequency and impact, are somewhat fewer than other reports. 69.6% of respondents identified interruption to the performance of clinical tasks as a potential negative impact of existing communication technologies.

Whilst relatively small in sample size, this study substantiated the findings of the earlier clinical governance-based analysis of the ICC framework at Austin Health. The high complexity and high variability in the performance of ICC, with an evolved, rather than designed, organisational technology stack, involving numerous communication methods, was confirmed by survey participants. Patient safety risks attributable through prior research to the mapped state of ICC practice, were confirmed to be present within our care environment.

Implicit in the simplification of ICC is the identification of the correct person with whom to communicate. Ideally, the allocation of specific and structurally stable clinical roles enables consistency of approach and process regardless of which person is allocated to that role in a given moment. This would theoretically reduce the delay in identification and contact with that role. Interestingly, the concept of ROLES at a higher and more generalised level suggests that different Roles may benefit from different communication tools. In this construct Innovator, Resource Investigator, Chair, Shaper, Evaluator, Team Worker, Organiser, Finisher (32) may have functional representations (in a health service sense) in roles such as Consultant, Registrar, Intern, Nurse, Administrative support, Nurse co-ordinator or Unit Manager. These functional roles may be present in all professional group teams or multi professional teams. Our findings suggest that each of these cohorts may have preferences for different tools and this has implications for design and function of any future enterprise system that purports to reduce the number of modalities in use.

The need for Austin Health to look beyond structured FCC processes, review ICC governance from a holistic system perspective, and consider communication technology as a fundamental component of clinical governance for patient care was confirmed by this study. Survey results also indicated that the existing clinical communication framework should be further reviewed as an environmental factor that could be negatively impacting upon staff role-satisfaction, wellbeing, and burnout. Experiential feedback provided by frontline clinicians including the level of use of each communication technology, as well as variations in technology use across different clinical disciplines provides critical insight to guide and prioritise future system improvements.

### Strengths of Study

Our study documents the perceived frequency and impact of multi-modality communications practices in a tertiary / quaternary public hospital in Victoria, Australia. It is broadly representative of many other Australian and International hospitals in the nature of the communications requirements and uses technology provided by international vendors for clinical communication. The literature supports our findings with respect to the impact of unstructured communications and multiple technology platforms on clinical inefficiency, interruptions and potential clinical errors. The survey respondents represented a wide variety of roles and levels of clinical responsibility, were self-selected to respond and were therefore motivated to share information they thought was important. The study confirms the range of clinical communication technology in use by frontline clinicians, validating the Health Service’s clinical governance based assessment of the existing communication framework. Our findings on the impact of existing clinical communication technologies in use, and the risk of high variation in clinical practice, are consistent with those reported in both Australian and international literature. The study forms an important baseline of current ICC practices in the context of the ever-increasing availability of new communication platforms and applications. We document the need to look beyond structured clinical handover to improve the safety of clinical communication practice and reduce the potential for communication associated patient harm. The impact of multi-modal ICT on interruption type and frequency is also observed. We also identify the predominantly role-based nature of ICC practice that requires technological change for greater efficiency. This study confirms the need to formalise ICC governance models and rationalise technology to reduce impacts and risks of multiple concurrent communication methods.

### Weaknesses of Study

The overall number of respondents was smaller than anticipated and the self-selection process may have inadvertently biased reporting. Reporting was subjective and there were no independent observational or time in motion validations of personal perceptions regarding the number of communications occurring in each respondent group, nor the impact on individual respondents. Although the communications systems in use are provided by multinational corporations and are part of international product suites, other hospitals may use them differently than in our hospital. Hospital cultures and communications norms vary across organisations and countries and so may not be representative in all hospitals. The rapid implementation of MS Teams (around the time of the COVID-19 pandemic) without clear guidance regarding use for clinical care or ‘rules of engagement’ in place may have biased the experience of this platform for some clinicians. Staff may have over or underestimated the potential time savings they could achieve if they were able to contact the right person on first attempt.

### Conclusions

Improving clinical communication safety and efficiency requires more attention to ICC practice and the technology available to suit current-day communication requirements.

Current systems involve multiple historic communication methods that evolved over time, were layered on top of each other and which enable variations in practice including personal preferences to occur. Inefficiency, delay, clinician frustration, paucity of documentation, incompleteness of information content and unnecessary interruption to clinical care are key deficiencies of the current communication systems.

This study details the clinician experience with the use of multiple ICC platforms and confirms the international experience of users in a multi-modality complex communications environment. The survey findings validated our prior clinical governance assessment of ICC practice and risks to patient safety.

Technical and organisational Areas identified to improve the quality and safety of ICC:

The use of enhanced and simplified technology for a single platform communication network.

Allocation of Key Organisational Roles to reduce dependency on rosters and lists to enable better “find first time” performance.

Recognition of individual role needs and preferences must be considered in future design.

Significant change management and modifications of work practice for some roles may be required if older technologies are to be removed from practice.

All such major changes to system behaviour must be evaluated for impacts on work efficiency, staff satisfaction and patient outcomes.

This study contributed to the design and user requirements specification for a simplified, single platform enterprise communications system “Baret Role Based Communicator” using MS Teams that was implemented during 2022-23 and which will be published elsewhere.

## Data Availability

All data produced in the present study are available upon reasonable request to the authors

## Acknowledgements

We are grateful for the support of Mr Alan Pritchard, Director of Information Technology, Mr Ray Van Kuyk, Exec Director Infrastructure and Planning and the hospital Executive for support in this and subsequent stages of the project culminating in the BARET Role Based Communications Application. We also thank Prof Kathleen Gray and Dr Hasan Ferdous of the Centre for Digital Transformation of Health for reviewing our draft.

## Appendix 1 : Pre-implementation Survey Instrument

### Pre-Implementation Survey Tool

1. What department / unit are you from?

- General Medicine
- Renal
- Emergency Department
- Radiology
- Level 7 Ward Nursing
- Intensive Care
- Bed Management

2. What role are you working in?

- h. Consultant
- Registrar
- j. Radiographer
- k. NUM
- l. ANUM
- m. Bedside nurse
- n. Intern / HMO
- o. Bed management / access
- p. Liaison / Coordinator
- q. Manager

3. What methods do you currently use to contact clinicians regarding clinical requests for information or tasks to be completed?

*Tick all that apply*

1. LAN page
2. Mobile phone
3. Landline phone
4. Cerner
5. Through switchboard
6. Messaging - text message
7. Messaging - What’s App
8. Teams
9. Email

4. How often do you use the following clinical communication methods?

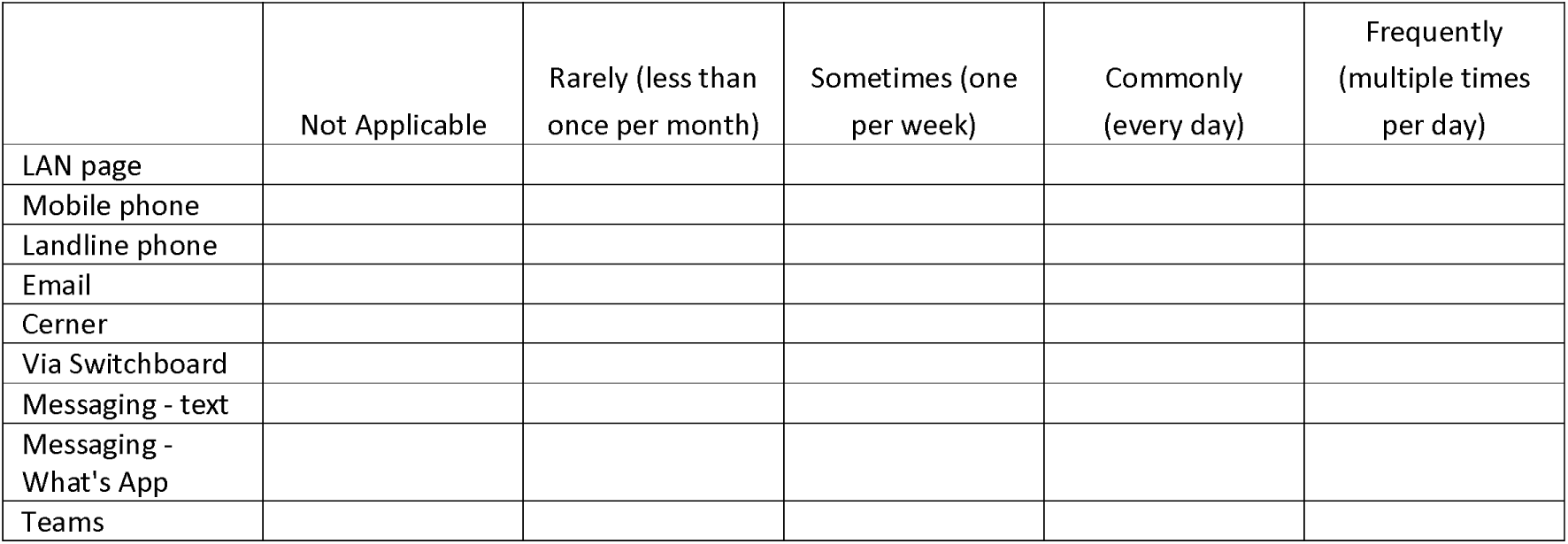

5. What is the timeliness of response for the following clinical communication methods?

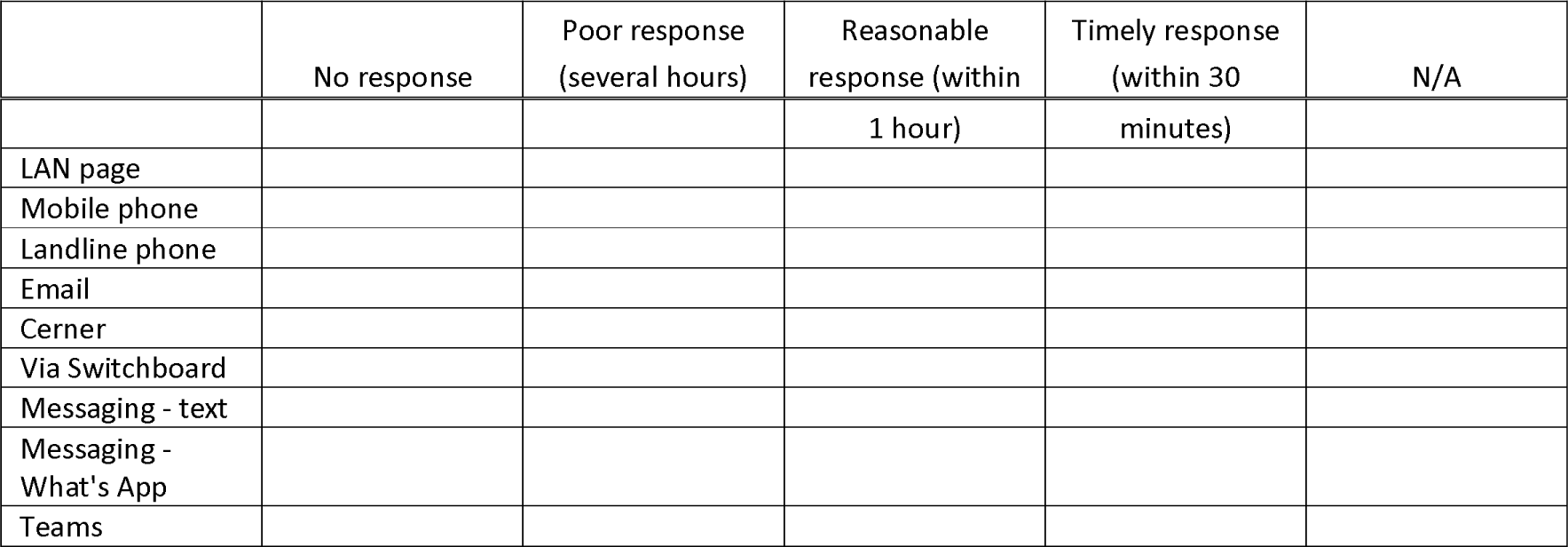

6. On average, how many times per shift do you need to make more than one attempt to locate the person required?

- Frequently (75-100% of the time)
- Commonly (50-75% of the time)
- Sometimes (10-50% of the time)
- Rarely (<10% of the time)
- Never

7. For each of the following roles, please indicate how many times per day (on average) you would initiate a communication.

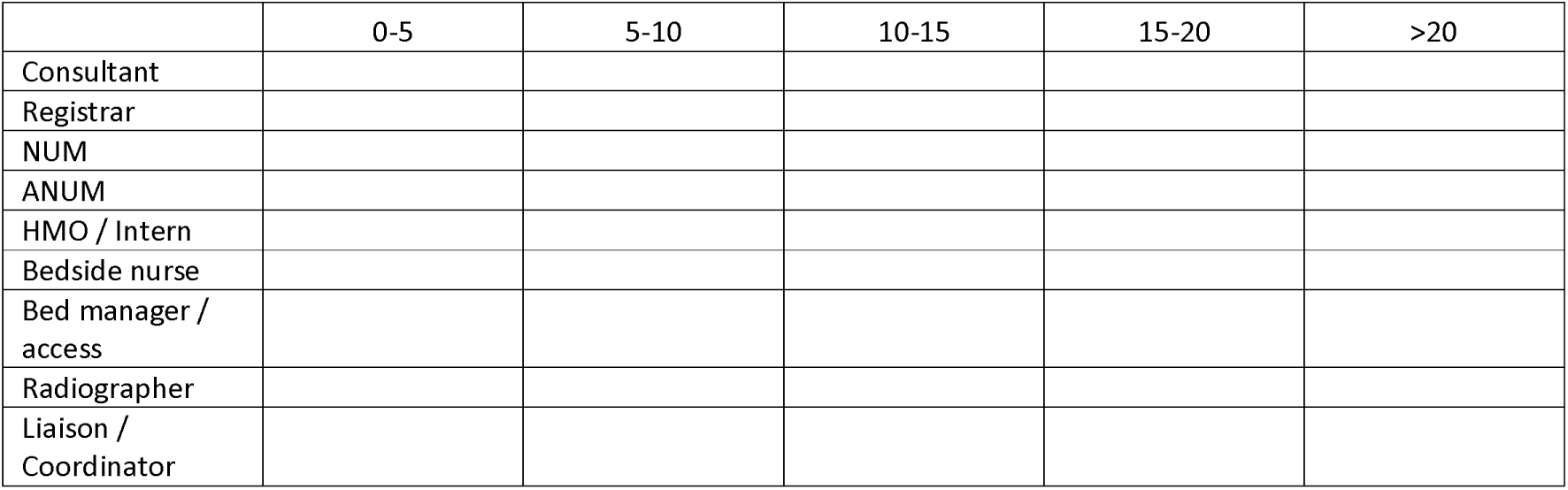

8. For each of the following roles, on average please rate the timeliness of response:

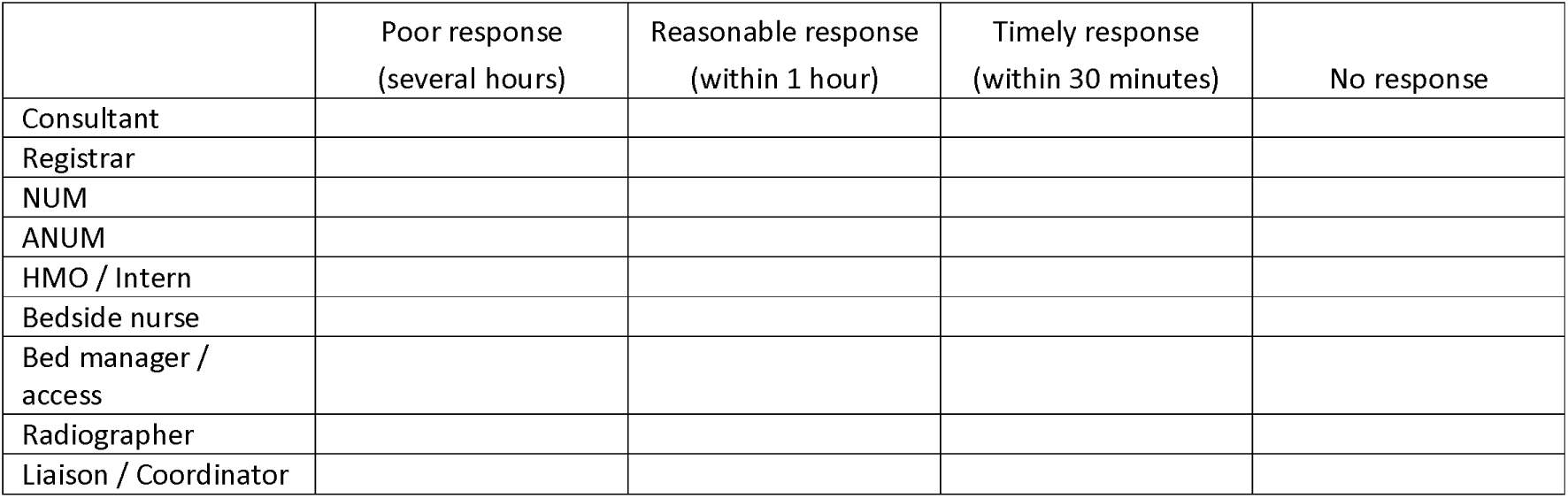

9. If you experience difficulties getting in contact with the right person, does this differ depending on the time of day, or day of the week?

1. Worse in AM
2. Worse in PM
3. Worse on weekends
4. No difference

10. On average, how much time per day do you think you would save if you were able to get in contact with the right clinician on the first attempt?

- < 15 minutes
- 15-30 minutes
- 30-60 minutes
- > 60 minutes

11. For existing communication methods, which of the following information do you receive consistently (eg. >80% of the time)?

Tick all that apply

- Patient identifiers
- Nature of the problem
- Call back / return information
- Who you are speaking / communicating with?
- Acknowledgement of your request

12. Do current formats of clinical communication provide documentation of the communication that occurred?

1. Yes
2. No
3. Not sure

13. How often would you be the incorrect recipient of a clinical request?

- Never
- Rarely (once per month)
- Sometimes (once per week)
- Usually (once per day)
- Often (multiple times per day)

14. What do you see as the potential risks with the multiple formats of clinical communication currently available?

Select all that apply.

1. Keeping track of communication and related actions
2. Missing communications
3. No response to communication
4. No documentation of communication occurring
5. 3 points of patient identification not used
6. Adverse patient event related to poor communication
7. Interruption to the performance of clinical tasks

15. Do current methods of clinical communication disrupt the performance of clinical patient care tasks and if so, how many times per shift does this occur?

- 2. 20+ per shift
- 10-20 per shift
- 5-10 per shift
- 0-5 per shift
- No interruptions

16. Any other comments?

### Ancillary On – Line data

#### Online Table 1

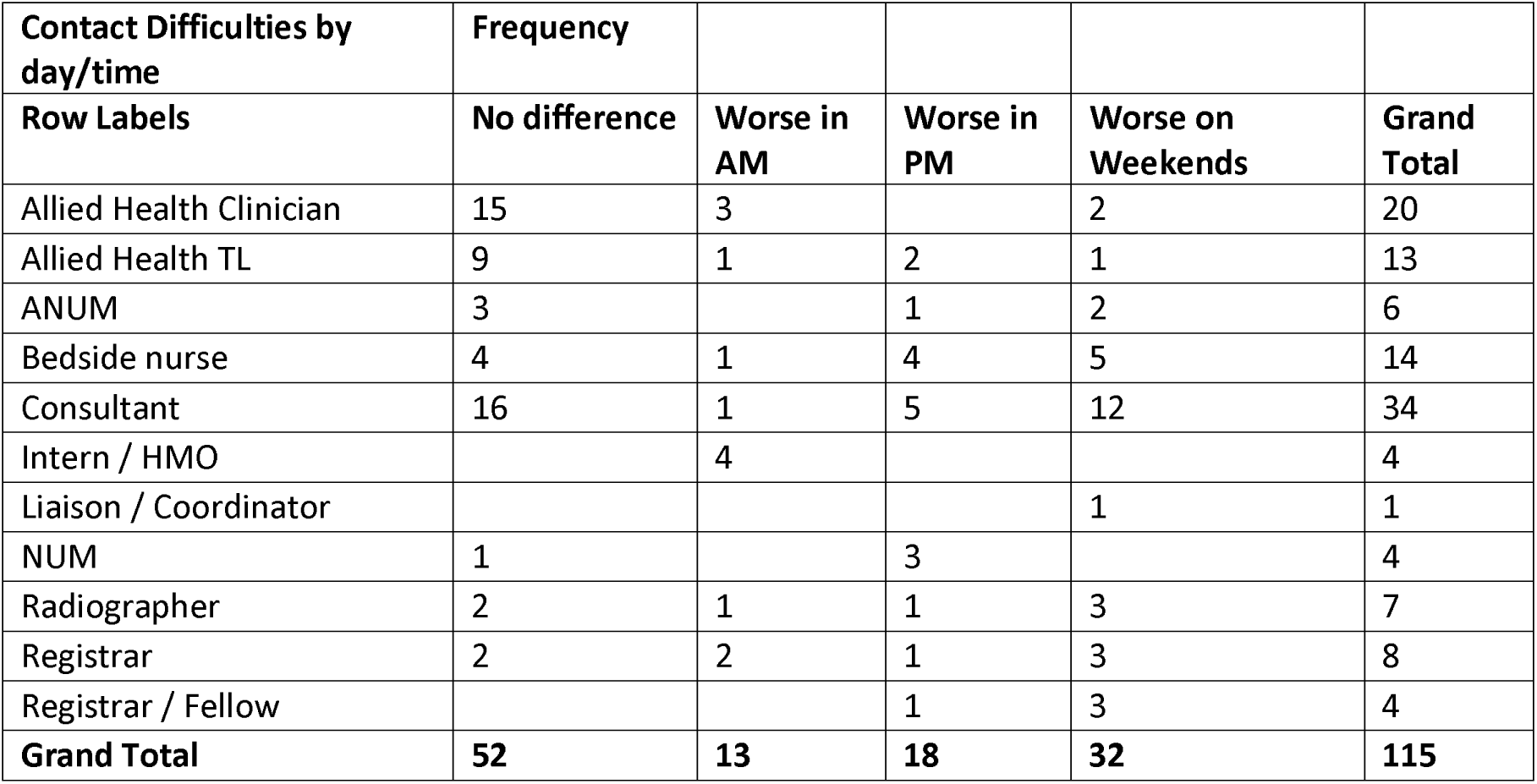

#### Online Table 2

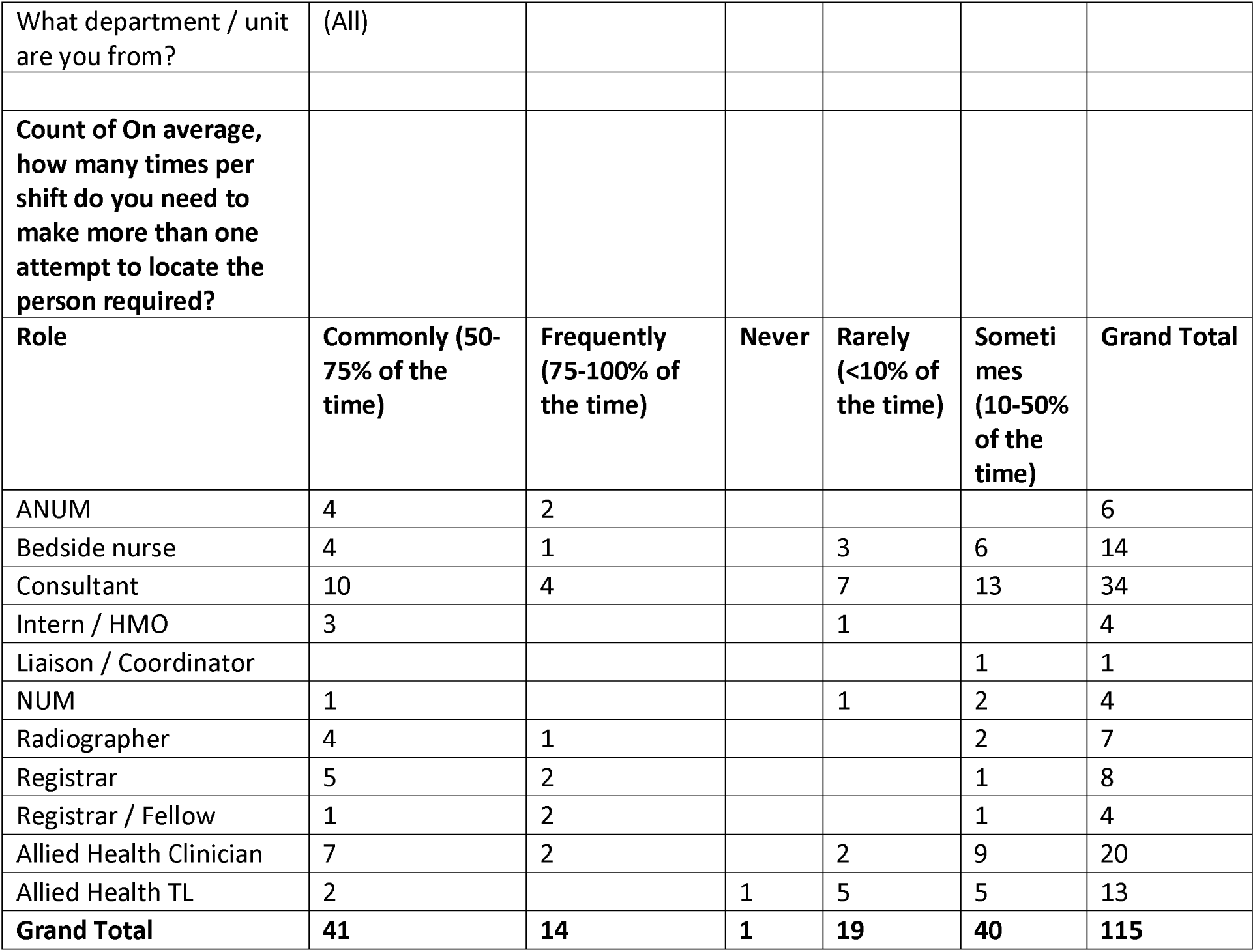

#### Online Table 3

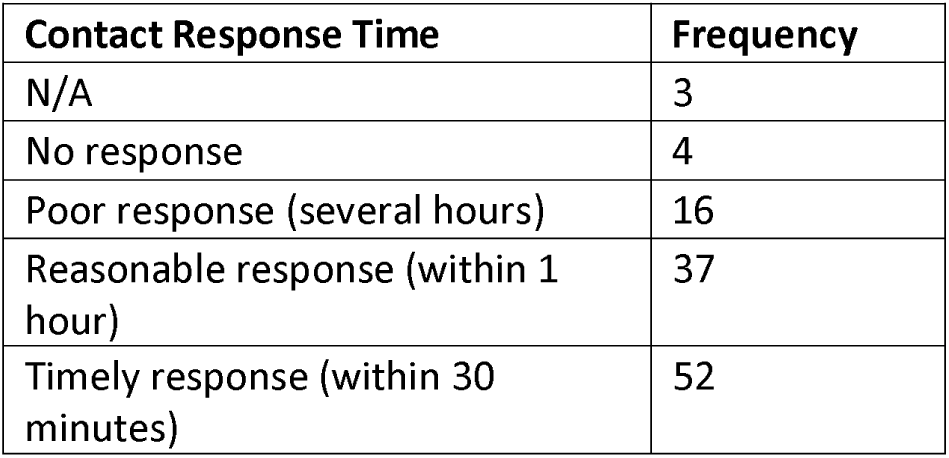

#### Online Table 4

Respondents were offered a list of possible impacts from current communication methods. They agreed with the options as follows.

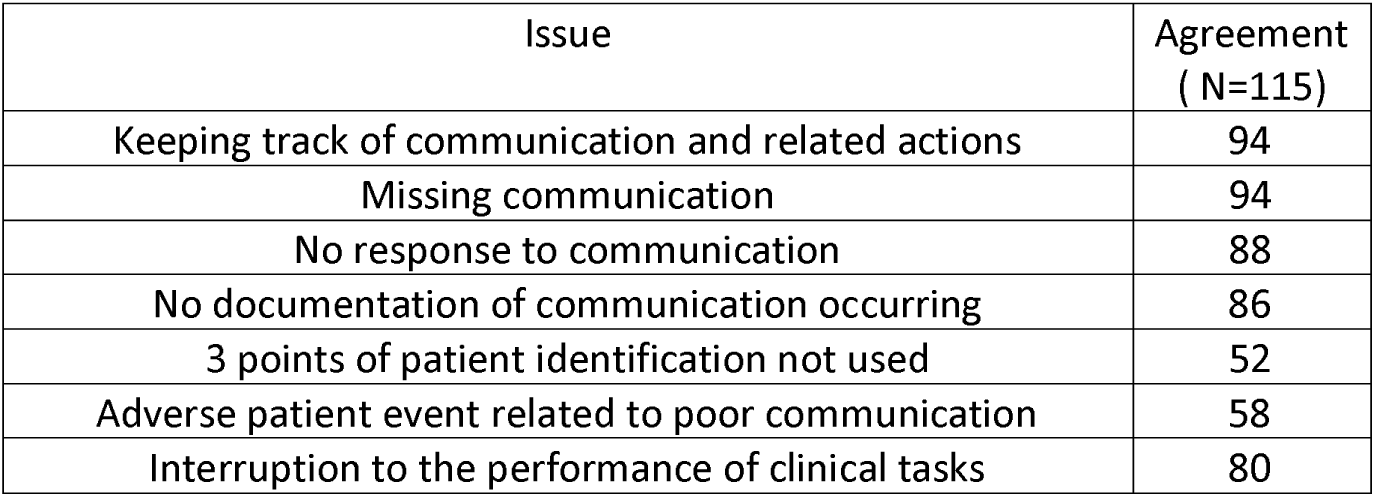

#### Online Table 5

Respondents endorsed the Problems and issues with current communications methodologies as follows. Only 20 % (approx.) of respondents agreed that they received important information more than 80% of the time.

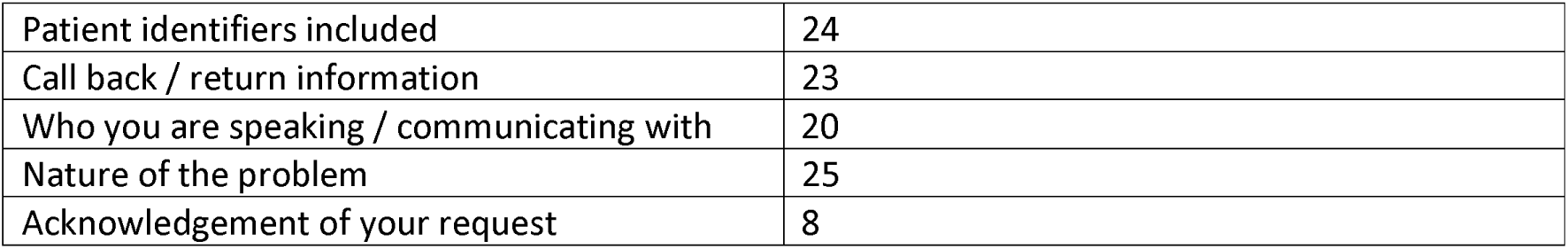

#### Online Fig 1

Respondent reported use of communications technologies

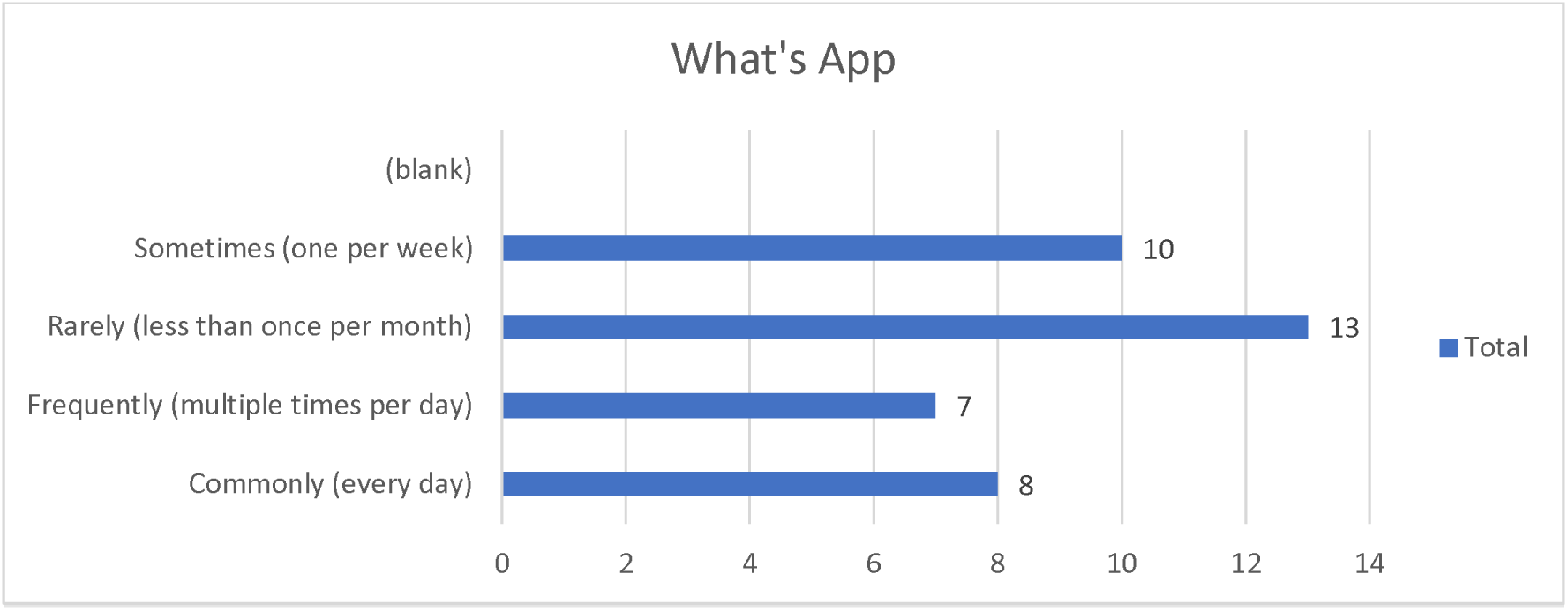

#### Online Table 6

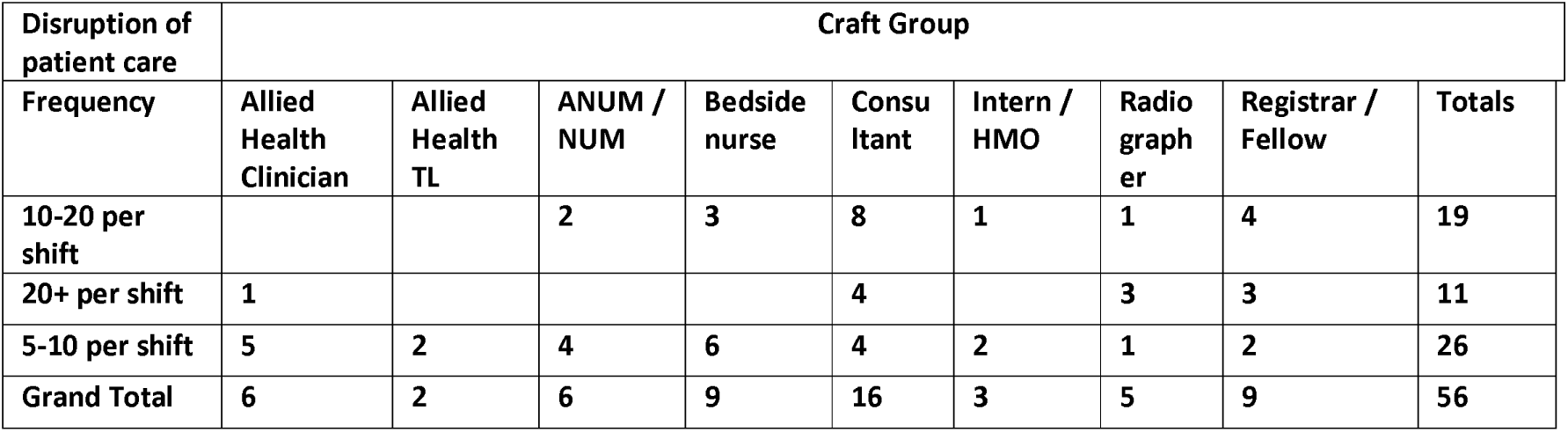

#### Online Fig 2

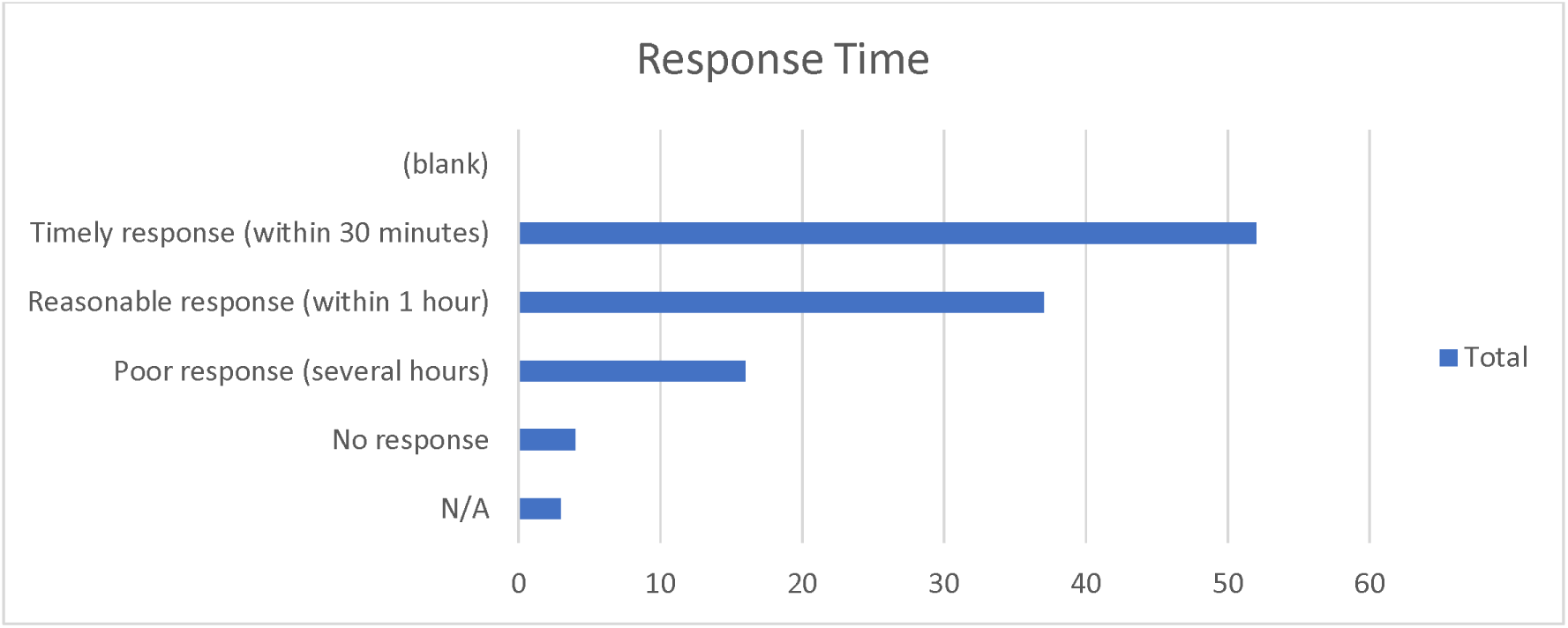

#### Online Table 7

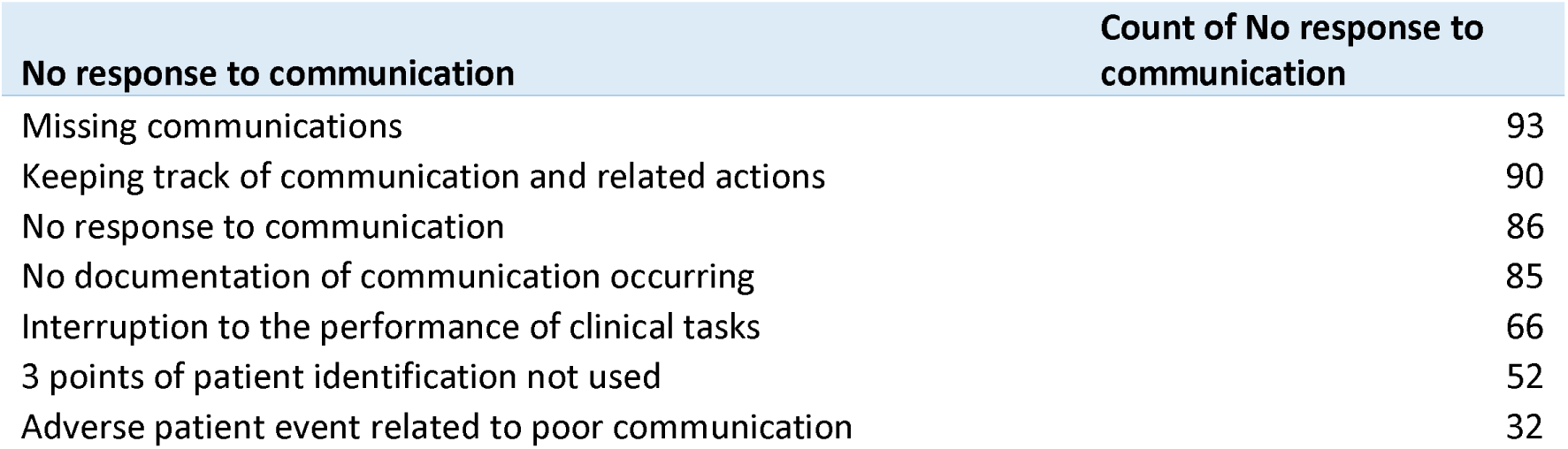

