## Supplemental information for "Technology Based Challenges of Informal Clinical Communication in an Australian tertiary referral hospital – A mixed methods assessment of The Need for Change"

Appendix 1 : Pre-implementation Survey Instrument

### Pre-Implementation Survey Tool

1.What department / unit are you from?

- General Medicine
- Renal
- Emergency Department
- Radiology
- Level 7 Ward Nursing
- Intensive Care
- Bed Management

2.What role are you working in?

- Consultant
- Registrar
- Radiographer
- NUM
- ANUM
- Bedside nurse
- Intern / HMO
- Bed management / access
- Liaison / Coordinator
- Manager

3.What methods do you currently use to contact clinicians regarding clinical requests for information or tasks to be completed?

*Tick all that apply*

1. LAN page
2. Mobile phone
3. Landline phone
4. Cerner
5. Through switchboard
6. Messaging - text message
7. Messaging - What's App
8. Teams
9. Email

4.How often do you use the following clinical communication methods?

|  | Not Applicable | Rarely (less than once per month) | Sometimes (one per week) | Commonly (every day) | Frequently (multiple times per day) |
| --- | --- | --- | --- | --- | --- |
| LAN page |  |  |  |  |  |
| Mobile phone |  |  |  |  |  |
| Landline phone |  |  |  |  |  |
| Email |  |  |  |  |  |
| Cerner |  |  |  |  |  |
| Via Switchboard |  |  |  |  |  |
| Messaging - text |  |  |  |  |  |
| Messaging - What's App |  |  |  |  |  |
| Teams |  |  |  |  |  |

5.What is the timeliness of response for the following clinical communication methods?

|  | No response | Poor response (several hours) | Reasonable response (within 1 hour) | Timely response (within 30 minutes) | N/A |
| --- | --- | --- | --- | --- | --- |
| LAN page |  |  |  |  |  |
| Mobile phone |  |  |  |  |  |
| Landline phone |  |  |  |  |  |
| Email |  |  |  |  |  |
| Cerner |  |  |  |  |  |
| Via Switchboard |  |  |  |  |  |
| Messaging - text |  |  |  |  |  |
| Messaging - What's App |  |  |  |  |  |
| Teams |  |  |  |  |  |

6.On average, how many times per shift do you need to make more than one attempt to locate the person required?

- Frequently (75-100% of the time)
- Commonly (50-75% of the time)
- Sometimes (10-50% of the time)
- Rarely (<10% of the time)
- Never

7.For each of the following roles, please indicate how many times per day (on average) you would initiate a communication.

|  | 0-5 | 5-10 | 10-15 | 15-20 | >20 |
| --- | --- | --- | --- | --- | --- |
| Consultant |  |  |  |  |  |
| Registrar |  |  |  |  |  |
| NUM |  |  |  |  |  |
| ANUM |  |  |  |  |  |
| HMO / Intern |  |  |  |  |  |
| Bedside nurse |  |  |  |  |  |
| Bed manager / access |  |  |  |  |  |
| Radiographer |  |  |  |  |  |
| Liaison / Coordinator |  |  |  |  |  |

8.For each of the following roles, on average please rate the timeliness of response:

|  | Poor response (several hours) | Reasonable response (within 1 hour) | Timely response (within 30 minutes) | No response |
| --- | --- | --- | --- | --- |
| Consultant |  |  |  |  |
| Registrar |  |  |  |  |
| NUM |  |  |  |  |
| ANUM |  |  |  |  |
| HMO / Intern |  |  |  |  |
| Bedside nurse |  |  |  |  |
| Bed manager / access |  |  |  |  |
| Radiographer |  |  |  |  |
| Liaison / Coordinator |  |  |  |  |

9.If you experience difficulties getting in contact with the right person, does this differ depending on the time of day, or day of the week?

1. Worse in AM
2. Worse in PM
3. Worse on weekends
4. No difference

10.On average, how much time per day do you think you would save if you were able to get in contact with the right clinician on the first attempt?

- < 15 minutes
- 15-30 minutes
- 30-60 minutes
- > 60 minutes

11.For existing communication methods, which of the following information do you receive consistently (eg. >80% of the time)?

Tick all that apply

- Patient identifiers
- Nature of the problem
- Call back / return information
- Who you are speaking / communicating with?
- Acknowledgement of your request

12.Do current formats of clinical communication provide documentation of the communication that occurred?

1. Yes
2. No
3. Not sure

13.How often would you be the incorrect recipient of a clinical request?

- Never
- Rarely (once per month)
- Sometimes (once per week)
- Usually (once per day)
- Often (multiple times per day)

14.What do you see as the potential risks with the multiple formats of clinical communication currently available?

Select all that apply.

1. Keeping track of communication and related actions
2. Missing communications
3. No response to communication
4. No documentation of communication occurring
5. 3 points of patient identification not used
6. Adverse patient event related to poor communication
7. Interruption to the performance of clinical tasks

15.Do current methods of clinical communication disrupt the performance of clinical patient care tasks and if so, how many times per shift does this occur?

- 20+ per shift
- 10-20 per shift
- 5-10 per shift
- 0-5 per shift
- No interruptions

16.Any other comments?

**Ancillary On – Line data**

**Online Table 1**

| **Contact Difficulties by day/time** | **Frequency** |  |  |  |  |
| --- | --- | --- | --- | --- | --- |
| **Row Labels** | **No difference** | **Worse in AM** | **Worse in PM** | **Worse on Weekends** | **Grand Total** |
| Allied Health Clinician | 15 | 3 |  | 2 | 20 |
| Allied Health TL | 9 | 1 | 2 | 1 | 13 |
| ANUM | 3 |  | 1 | 2 | 6 |
| Bedside nurse | 4 | 1 | 4 | 5 | 14 |
| Consultant | 16 | 1 | 5 | 12 | 34 |
| Intern / HMO |  | 4 |  |  | 4 |
| Liaison / Coordinator |  |  |  | 1 | 1 |
| NUM | 1 |  | 3 |  | 4 |
| Radiographer | 2 | 1 | 1 | 3 | 7 |
| Registrar | 2 | 2 | 1 | 3 | 8 |
| Registrar / Fellow |  |  | 1 | 3 | 4 |
| **Grand Total** | **52** | **13** | **18** | **32** | **115** |

**Online Table 2**

| What department / unit are you from? | (All) |  |  |  |  |  |
| --- | --- | --- | --- | --- | --- | --- |
| **Count of On average, how many times per shift do you need to make more than one attempt to locate the person required?** |  |  |  |  |  |  |
| **Role** | **Commonly (50-75% of the time)** | **Frequently (75-100% of the time)** | **Never** | **Rarely (<10% of the time)** | **Sometimes (10-50% of the time)** | **Grand Total** |
| ANUM | 4 | 2 |  |  |  | 6 |
| Bedside nurse | 4 | 1 |  | 3 | 6 | 14 |
| Consultant | 10 | 4 |  | 7 | 13 | 34 |
| Intern / HMO | 3 |  |  | 1 |  | 4 |
| Liaison / Coordinator |  |  |  |  | 1 | 1 |
| NUM | 1 |  |  | 1 | 2 | 4 |
| Radiographer | 4 | 1 |  |  | 2 | 7 |
| Registrar | 5 | 2 |  |  | 1 | 8 |
| Registrar / Fellow | 1 | 2 |  |  | 1 | 4 |
| Allied Health Clinician | 7 | 2 |  | 2 | 9 | 20 |
| Allied Health TL | 2 |  | 1 | 5 | 5 | 13 |
| **Grand Total** | **41** | **14** | **1** | **19** | **40** | **115** |

**Online Table 3**

| **Contact Response Time** | **Frequency** |
| --- | --- |
| N/A | 3 |
| No response | 4 |
| Poor response (several hours) | 16 |
| Reasonable response (within 1 hour) | 37 |
| Timely response (within 30 minutes) | 52 |

**Online Table 4**

Respondents were offered a list of possible impacts from current communication methods. They agreed with the options as follows.

| Issue | Agreement ( N=115) |
| --- | --- |
| Keeping track of communication and related actions | 94 |
| Missing communication | 94 |
| No response to communication | 88 |
| No documentation of communication occurring | 86 |
| 3 points of patient identification not used | 52 |
| Adverse patient event related to poor communication | 58 |
| Interruption to the performance of clinical tasks | 80 |

**Online Table 5**

Respondents endorsed the Problems and issues with current communications methodologies as follows. Only 20 % (approx.) of respondents agreed that they received important information more than 80% of the time.

| Patient identifiers included | 24 |
| --- | --- |
| Call back / return information | 23 |
| Who you are speaking / communicating with | 20 |
| Nature of the problem | 25 |
| Acknowledgement of your request | 8 |

**Online Fig 1**

Respondent reported use of communications technologies

What's App

|  | Total |
| --- | --- |
| Commonly (every day) | 8 |
| Frequently (multiple times per day) | 7 |
| Rarely (less than once per month) | 13 |
| Sometimes (one per week) | 10 |

**Online Table 6**

| **Disruption of patient care** | **Craft Group** | | | | | | | | |
| --- | --- | --- | --- | --- | --- | --- | --- | --- | --- |
| **Frequency** | **Allied Health Clinician** | **Allied Health TL** | **ANUM / NUM** | **Bedside nurse** | **Consultant** | **Intern / HMO** | **Radiographer** | **Registrar / Fellow** | **Totals** |
| **10-20 per shift** |  |  | **2** | **3** | **8** | **1** | **1** | **4** | **19** |
| **20+ per shift** | **1** |  |  |  | **4** |  | **3** | **3** | **11** |
| **5-10 per shift** | **5** | **2** | **4** | **6** | **4** | **2** | **1** | **2** | **26** |
| **Grand Total** | **6** | **2** | **6** | **9** | **16** | **3** | **5** | **9** | **56** |

| **Online Fig 2** |
| --- |

Response Time

|  | Total |
| --- | --- |
| N/A | 3 |
| No response | 4 |
| Poor response (several hours) | 16 |
| Reasonable response (within 1 hour) | 37 |
| Timely response (within 30 minutes) | 52 |

**Online Table 7**

| **No response to communication** | **Count of No response to communication** | |
| --- | --- | --- |
| Missing communications | | 93 |
| Keeping track of communication and related actions | | 90 |
| No response to communication | | 86 |
| No documentation of communication occurring | | 85 |
| Interruption to the performance of clinical tasks | | 66 |
| 3 points of patient identification not used | | 52 |
| Adverse patient event related to poor communication | | 32 |
